# Short and midterm outcomes of surgical repair of acute type A aortic dissection concomitant CABG vs. ECMO support: A retrospective study

**DOI:** 10.1101/2024.06.05.24308525

**Authors:** Dong Zhang, Gui-Jun Zhu, Ming-Jun Gao, Xiang-Yang Wei, Zhe Yan, Bin Li, Xing-Peng Chen, Xiao-Lin Wang, Yu-Sheng Shu

## Abstract

**Objective:** Intraoperative surgical repair of acute type A aortic dissection, sometimes we would encounter special cases that failed to wean from CPB. In this study, we aimed to retrospectively analyze that the indication, clinical experience and short and midterm outcomes of surgical repair of acute type A aortic dissection concomitant CABG or ECMO support in patients who failed to wean from CPB.

**Methods:** A total of 532 consecutive patients underwent emergent surgery for TAAD in a single institution between Jan 2018 and Jan 2023. And categorized into CABG group and ECMO group based on surgical approach. Preoperative, intraoperative and postoperative variables were assessed and analyzed. Outcomes of the patients were followed up until five years from discharge of hospital.

**Results:** Overall in-hospital mortality was determined to be 15.7% for CABG group and 73.3% for ECMO group (*P*=0.001). The operation time, CPB time, extracorporeal circulation assisted time, 24-hour traffic diversion in CABG group were less than ECMO group, and had statistically different between two groups (*P*=0.039, *P*=0.007, *P*<0.001, *P*<0.001). Higher morbidity of delayed chest closure, low cardiac output syndrome, and lower limb osteofascial compartment syndrome in the ECMO group than the CABG group, but not statistically significant (*P*=0.139, *P*=1, *P*=0.524).

5-years follow-up mortality after discharge had no statistically different between two groups (*P*=1).

**Conclusion:** For the patients who failed to wean from CPB, surgical repair of acute type A aortic dissection Concomitant CABG can provide more excellent short and midterm outcomes than ECMO support. However, concomitant CABG are also associated with long-term complications of the great saphenous vein embolization and severe tricuspid valve regurgitation.

## 1. Introduction

TAAD is one of the most urgent surgical emergencies in cardiac surgical patients. Despite significant improvements in surgical techniques, cardiopulmonary bypass (CPB) practices, cerebral protection procedures, and perioperative management, data published in the International Registry of Acute Aortic Dissection (IRAD) show that the mortality rate in TAAD surgery is still approximately 20% [1]. In general, most patients have successful surgery and a good prognosis, sometimes some patients failed to wean from cardiopulmonary bypass due to several reasons, such as intra-coronary gas embolism, electrolyte disturbance, low blood sugar, CAS, acute myocardial infarction, severe myocardial edema, and so on. If the conventional adjustment of vasoactive drugs fails to achieve the desired therapeutic effect, what should we do next? The Cardiac massage, correcting electrolyte disturbance, increasing coronary perfusion pressure, delayed chest closing, continuing the cardiopulmonary bypass, CABG, ECMO support?

Recently, there are few reports about what to do when come to these special cases that failed to wean from CPB. If that, CABG or ECMO? Intraoperative selection of appropriate personalized adjuncts is crucial. Our center has managed 34 cases of type A aortic dissection which are failed to wean from CPB over the past five years. The aim of this study is to clarify the intraoperative decision making for the patients who failed to wean from CPB, indications of CABG or ECMO and significant clinical experience, the short and midterm outcomes of surgical repair of acute type A aortic dissection concomitant CABG vs. ECMO support in a high-volume single-center experience.

## 2. Patients and methods

This is a retrospective analysis of prospectively collected data of patients who underwent emergency surgery for TAAD at our institution. A total of 532 consecutive patients underwent emergent surgery for TAAD in a single institution between Jan 2018 and Jan 2023. In all patients, the aortic dissection was diagnosed with the aid of either computerized tomography before surgical treatment. Clinical variables including basic characteristics, perioperative data, in-hospital and follow-up outcome. Ultimately, 34 patients out of the 532 patients who had undergone surgical repair of acute type A aortic dissection concomitant CABG or ECMO support were enrolled in our study. Patients received surgical repair of acute type A aortic dissection concomitant CABG or ECMO support were 19 and 15, respectively. The preoperative characteristics were listed in Table 1. Follow-up data were obtained through clinical interviews and telephone, the follow-up time was 1.3-66.8 months, and the average follow-up time was 44.1±13.9 months.

**Table I.**
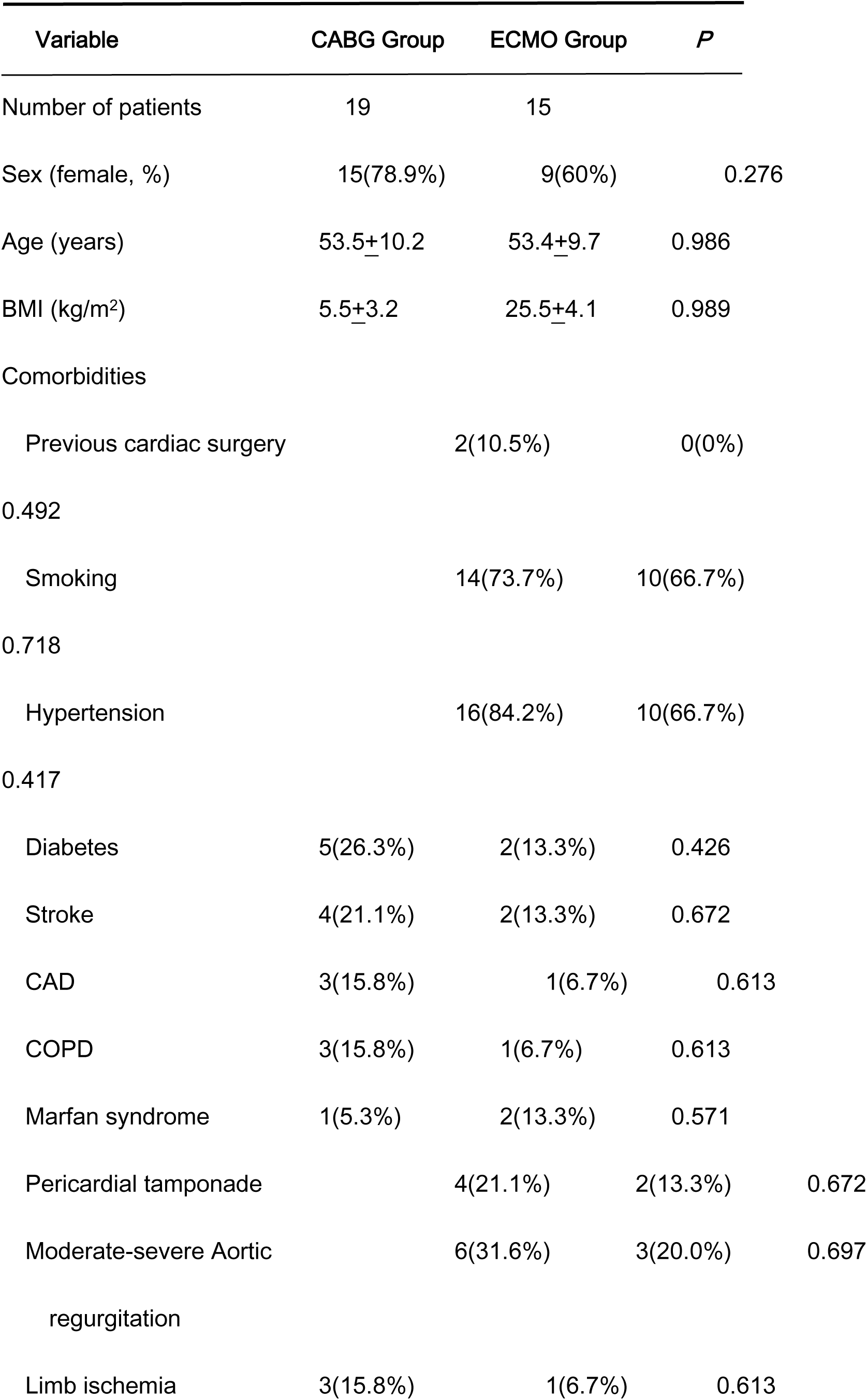
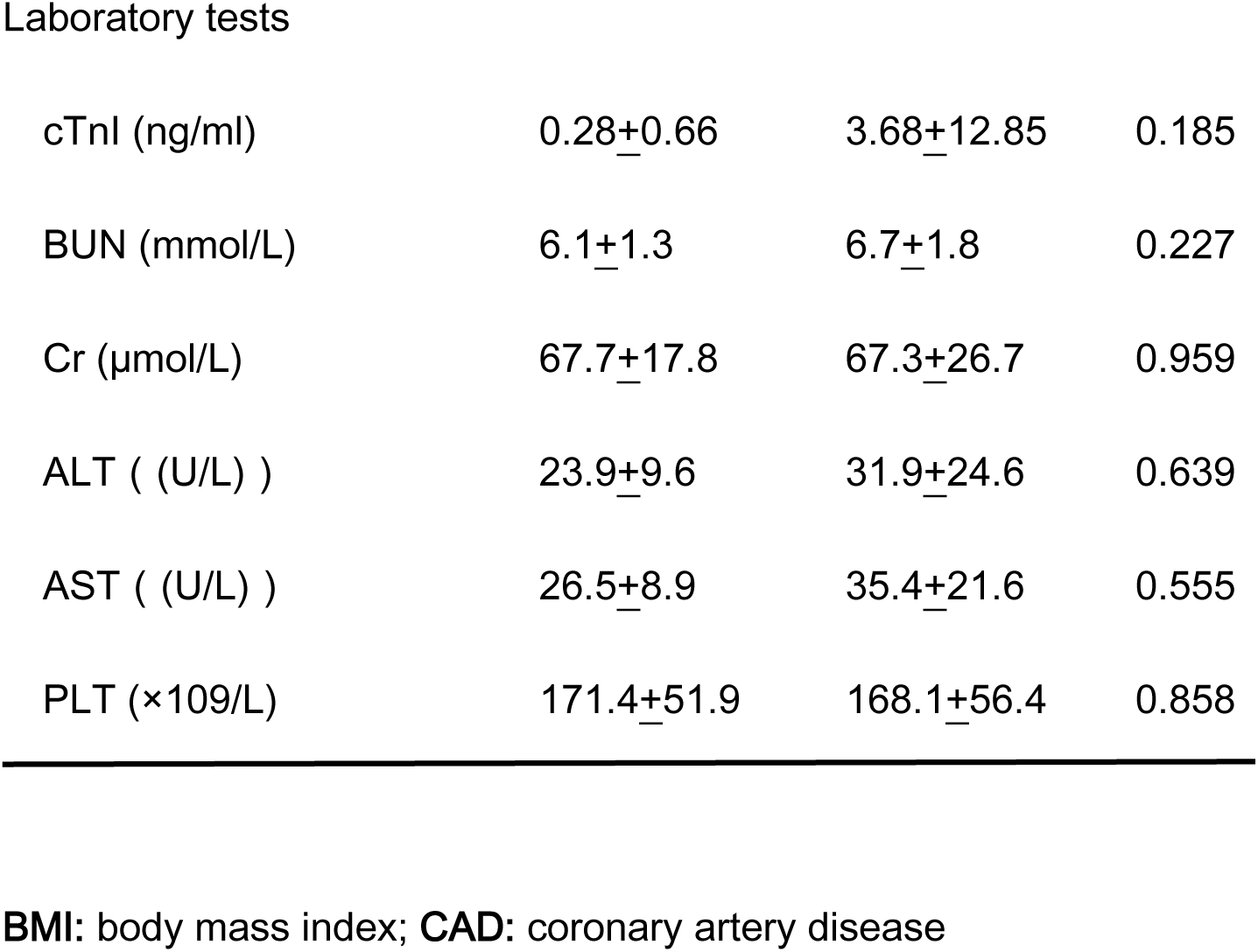
Demographics and Characteristics of Patients.

## 3. Surgical procedure

All patients were carried out by a median sternotomy after general anesthesia. We preferred to establish the cardiopulmonary bypass through innominate artery and right atrium, sometimes right axillary artery or femoral artery. The left heart was vented through the right superior pulmonary vein avoided the ventricular dilation during the operation. Once ventricular fibrillation was identified, a cross-clamp was placed across the distal ascending aorta, which was then incised longitudinally at the distal sinus tubular junction. Myocardial protection was achieved with a cold blood cardioplegic solution via the non-dissected coronary ostium, or retrograde, through the coronary sinus. When patients were concomitant with myocardial ischemia, coronary arteries bypass grafting was considered to perform using the saphenous vein. And an intermittent cold blood cardioplegic solution was then administered through the saphenous vein at a speed of 1-2r/s. Aortic root surgery was completed during the cooling process. In general, an artificial artery strip was used to reinforce the aortic root dissection circumferentially in the sinotubular junction level in a sandwiched manner. Circulatory arrest was initiated when the nasopharyngeal temperature reached 17°C-22°C, CPB was discontinued and adjusted the direction of the innominate artery intubation. The selective cerebral perfusion was performed through the innominate artery and left common carotid artery. Then replaced total arch and deployed frozen elephant trunk in descending aorta. The lower body circulation was reinstated after anastomosis of the distal aortic arch was complete. we reconstructed the left common carotid artery to achieve bilateral perfusion during the heart rewarming, then reconstructed the innominate and subclavian arteries, Finally, proximal anastomosis of the saphenous vein graft was completed. At this point, the main surgical steps were completed. If the ECMO support needed, the VA-ECMO was implanted immediately in the operation room or ICU through the femoral vein and artery, and we always used a 10 Fr cannula connected to the side port of the femoral cannula to avoid the lower limb ischemia. However, final surgical strategy was made by surgeons according to intraoperative findings.

## 4. Statistical analysis and follow-up

Categorical variables are represented as frequency distributions and single percentages. Values of continuous variables are expressed as mean ± standard deviation. Normally distributed continuous variables were compared using T-test, non-normally distributed continuous variables using Mann-Whitney U test, and categorical variables using χ2 and Fisher’s exact test. Long-term survival analysis was conducted using Kaplan-Meier method with log-rank test for group comparisons. All statistical tests were two-sided. Results were considered statistically significant at a level of *P*< 0.05. The entire analytical process was executed using IBM SPSS statistical software version 21.0.

Survived patients were followed yearly in our outpatient department or by phone to evaluate their clinical status.

## Results

Between Jan 2018 and Jan 2023, of 532 screened patients, 34 who met the inclusion criteria were assigned. Overall, 34 patients were followed up in both groups, with 19 patients underwent surgical repair of acute type A aortic dissection Concomitant CABG and 15 with ECMO support; all patients underwent complete protocol-guided follow-up. Patient selection and outcomes are illustrated in Figure 1.

**Fig 1.**
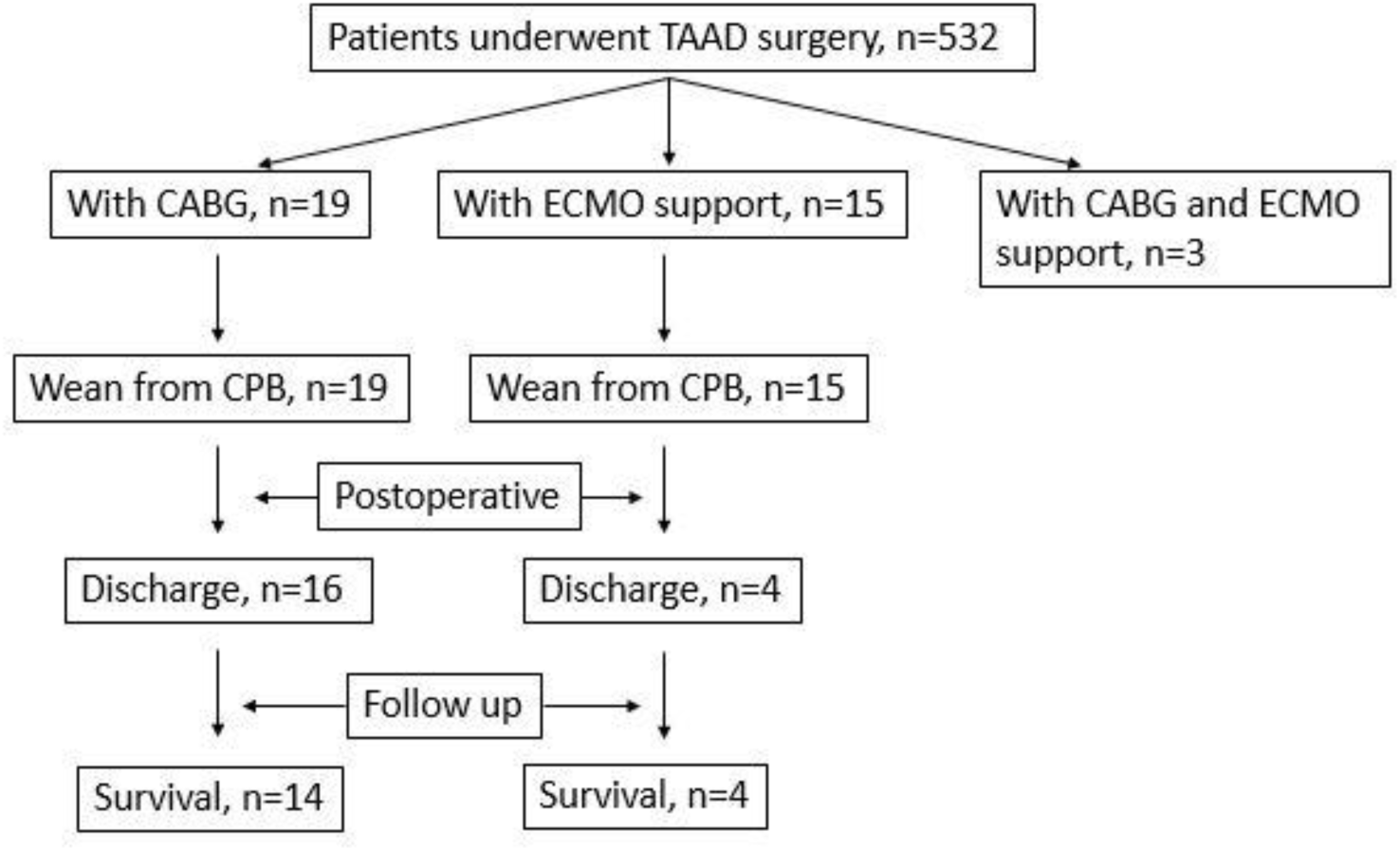
The enrollment, allocation, and follow-up of TAAD patients who underwent surgical repair of acute type A aortic dissection Concomitant CABG or ECMO support.

Baseline and demographic characteristics, comorbidity, laboratory tests, clinical manifestations of patients were showed in Table 1. Age, gender, and the presence of comorbidities, laboratory tests were not significantly different in all patients. Moreover, in the CABG group, cTnI values were generally higher than in the ECMO group, but not statistically significant (*P=*0.185). And patients with preoperatively diagnosed coronary artery disease who with PCI or pharmacologic therapy were 3 and 1, respectively, but not statistically significant (*P*>0.05).

Table 2 provides detailed procedural characteristics of two groups. The operation time, CPB time, extracorporeal circulation assisted time, 24-hour traffic diversion, in-hospital mortality in CABG group were less than ECMO group, and had statistically different between the two groups (*P*=0.039, *P*=0.007, *P*<0.001, *P*<0.001, *P*=0.001). There was no significant difference between the two groups for the ventilator-assisted time and ICU stay time (*P*=0.395, *P*=0.579), perhaps because most of the patients in the ECMO group died within a short period of time, which also greatly reduced them. Meanwhile, we also found the higher morbidity of delayed chest closure, low cardiac output syndrome, and lower limb osteofascial compartment syndrome in the ECMO group than the CABG group, but not statistically significant (*P*=0.139, *P*=1, *P*=0.524).

**Table 2.**
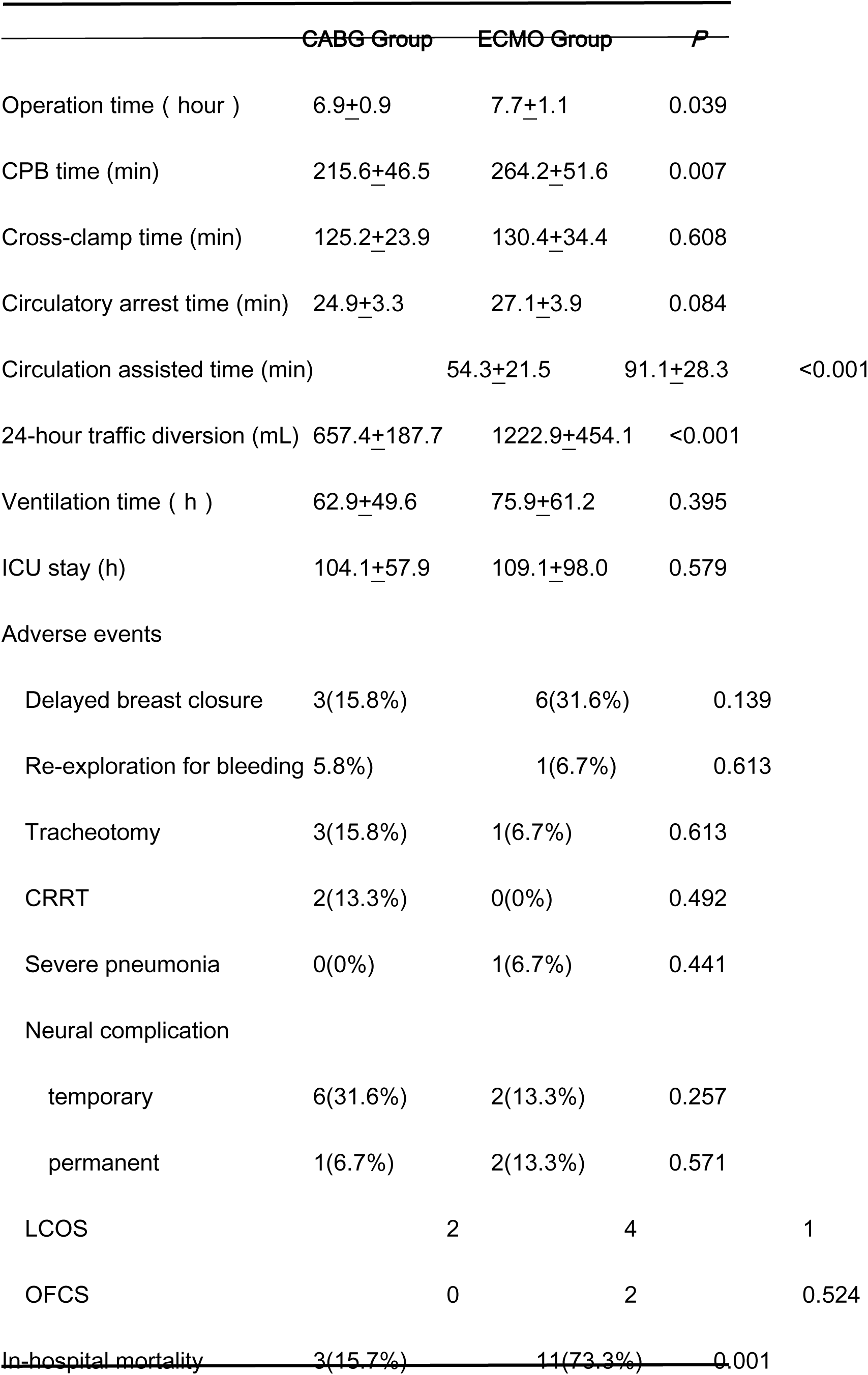
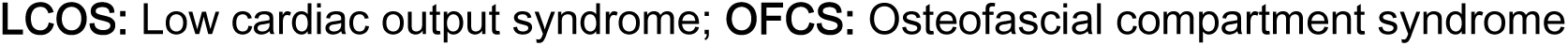
Procedural Characteristics and Outcomes before discharge.

Intraoperatively, we adopted the principle of prioritizing the aortic root. As soon as we found coronary artery involvement and require CABG, we immediately took the saphenous vein as a bypass vessel and performed myocardial arrest fluid perfusion through the saphenous vein. The Details of coronary artery bypass surgery and early patency were shown in Table 3. At follow-up, there were 2 right coronary bridge occlusions, 1 anterior descending bridge vessel occlusion, and 1 death, whereas the surviving patients of them perhaps survived because of the presence of collateral circulation or other vascular retrograde perfusion.

**Table 3.** Details of coronary artery bypass surgery and early patency.

| Location | Number | Early patency |
| --- | --- | --- |
| LAD | 3 | 0 |
| RCA | 12 | 11 |
| LAD+RCA | 4 | 1 |

Table 4 summarizes 5-years follow-up mortality and morbidity after discharge. Two patients died because of cardiac arrest due to bridge vessel occlusion and massive cerebral hemorrhage. we also found that the morbidity of moderate to severe tricuspid regurgitation in CABG group was higher than that ECMO group, but not statistically significant (*P*=0.569). The main reasons for secondary postoperative surgery in both groups were severe tricuspid regurgitation and tearing of the coronary artery opening, particularly the right coronary artery.

**Table 4.** 5-years follow-up Mortality and Morbidity after discharge.

|  | CABG Group | ECMO Group | *P* |
| --- | --- | --- | --- |
| Mortality | 2 | 0 | 1 |
| Middle-severe aortic<br>valve regurgitation | 0 | 0 |  |
| Middle-severe tricuspid<br>valve regurgitation | 6 | 1 | 0.569 |
| EF | 0.62±0.03 | 0.63±0.02 | 0.543 |
| Bridging vascular occlusion | 2 | 0 | 1 |
| Secondary interventions | 4 | 0 | 0.516 |
| Repair of coronary artery | 2 | 0 | 1 |
| <u>TVP</u> | <u>2</u> | <u>0</u> | <u>1</u> |

## Discussion

Owing to the advancements of diagnostic tools, surgical technique, and postoperative management, the outcome of aortic repair for acute type A aortic dissection (TAAD) has improved, and hospital mortality in patients with TAAD after aortic repair was decreased to 11% in a recent report [2]. For patients were failed to wean from cardiopulmonary bypass the mortality could be higher. In this study, we systematically summarized the characteristics, surgical strategies, and prognosis of patients with surgical repair of acute type A aortic dissection Concomitant CABG or ECMO support. For patients who failed to wean from cardiopulmonary bypass, we adopted intraoperative ECMO support and gauze tamponade for delayed closure of the chest previously, but the 24-hour postoperative drainage was still higher than the CABG group (*P*<0.001). On the one hand, the high depletion of preoperative coagulation factor, the fragile, dissected aorta and intraoperative hypothermia contribute to impaired postoperative hemostatic function [3], the other hand, ECMO anticoagulation could also increase bleeding. However, in-hospital mortality was significantly higher in the ECMO group than in the CABG group (*P* =0.001), Kai Zhang et al reported cross-clamp time and CPB time were identified as risk factors of operative mortality in the univariate analysis [4]. Most of these patients died within a short period of time because of postoperative low cardiac output syndrome, which explains not statistically significant in postoperative ventilator-assisted respiration time and ICU stay time between the ECMO group and CABG group (*P*=0.395, *P* =0.579). At present, when we encountered patients who failed to wean from cardiopulmonary bypass, we could not determine whether there were lesions of the coronary arteries due to routine preoperative coronary angiography or coronary CTA in AAAD patients is not performed at our Institution, so we need determine whether there is an indication for CABG. Although ECMO support has been widely employed in the treatment of heart and lung failure caused by many kinds of disease [5]. To some extent, it could reduce the burden on the heart, but perhaps it has not addressed the fundamental issue. Furthermore, CABG is a relatively routine and simple procedure and the great saphenous vein graft (SVG) is the main graft for CABG in STAAD because of its convenience and rapid access. Although individual patients were unable to wean from cardiopulmonary bypass and required ECMO support, postoperative mortality was significantly lower in the CABG group than ECMO group (n=15.7%, n =73.3%, *P*=0.001). The predominance of right coronary bypass grafts in these patients may be related to the anatomical relationship that right coronary locates at the anterior wall of the ascending aorta, bears a higher shear force from the blood flow, and no support from the pulmonary artery.

The postoperative follow-up of patients in both groups, for the patients having long-term survival, cardiac function returned to almost normal. And no severe coronary stenosis was found in ECMO group by coronary CTA or coronary angiography. However, we found two problems: First, preoperative patients had less-moderate tricuspid regurgitation, but there was a higher incidence of moderate-severe tricuspid regurgitation in the CABG group during the follow-up period. This may be attributed to two reasons: 1. Influenced by the anatomical location of the right coronary artery encircling the tricuspid valve, and acute ischemia of the tricuspid papillary muscle due to preoperative right coronary involvement. 2. Increased right heart burden due to cabrol shunt. Therefore, we recommend concomitant tricuspid valvuloplasty for preoperative patients with moderate-severe tricuspid regurgitation, especially those with right coronary artery involvement, in order to avoid secondary surgery in the future. Second, although postoperative antiplatelet therapy was given in the CABG group, some patients were found to have occlusion of the bridge vessel within 5 years after bypass surgery, which may be related to the use of the great saphenous vein graft as the bridge vessel and the proximal anastomotic location, and the fact that the distal anastomosis location of the bridge vessel is usually located at the left and right coronary trunks, so in the long term, it may be better to apply the internal mammary artery. Some studies report the use of LITA-LAD with a good patency rate in this setting [6].

The main manifestations of failing to wean from cardiopulmonary bypass are single or double ventricular pulse weakness, low blood pressure, high pulmonary artery pressure and central venous pressure. According to our 5-year experience, the main reasons for failing to wean from cardiopulmonary bypass are: 1. Low cardiac output syndrome, the most common, and which has three causes: (1) Severe myocardial ischemia caused by the coronary artery involved and ischemia-reperfusion injury; (2) Inadequate intraoperative myocardial protection; (3) Long-term chronic coronary atherosclerosis. 2. Severe myocardial edema, for myocardial edema, we often take delayed closure of the chest or open both sides of the pleura, as well as incise pericardium parallelly to the phrenic nerve to give the heart a large enough space to beat. If that’s not possible, we could trim the ends of two 20 ml syringes into gear-like grooves, prop the sternum, sutured the skin.3. the left heart failure caused by long-term aortic regurgitation (Marfan syndrome, Aortic root aneurysm, Congenital diplastic malformation).

For these patients who require CABG or ECMO support, accurate intraoperative indications are critical. Indications for CABG: (1) Weakness of single or double ventricle beat; (2) Severe involvement of the coronary arteries, especially Neri type B and C; (3) Severe stenotic lesions of the coronary arteries themselves or intraoperative detection of significant atheromatous plaques; (4) varying epicardium temperatures or purplish color (presenting like myocardial ischemia), and electrocardiographic ST-segment changes. Indications for ECMO: (1) High dosage of cardiovascular active drugs, severe myocardial edema with cardiac incompetence; (2) Severe coronary artery involvement or other reasons that unable to perform CABG; (3) Unexplained circulatory and respiratory failure.

In terms of the postoperative management of surgical repair of acute type A aortic dissection concomitant CABG or ECMO support, the main concern is the prevention and management of complications. Postoperative management of patients with CABG are mainly the prevention of bleeding (gastrointestinal emergence and cerebral hemorrhage) and bridge vessel occlusion associated with antiplatelet therapy. For patients with very high troponin values, intraoperative findings of coronary artery involvement have been severe and require combined CABG. Although facing the risk of myocardial ischemia-reperfusion injury and ECMO support, these patients tend to have a bad aortic root and higher risk of death, so we don’t wait until there is a tendency for troponin to decrease, and often operate early to improve myocardial ischemia, and most patients with high cTnI recover well after surgery. In contrast, there are many complications for patients with ECMO support.

The main complication is hemorrhage (gastrointestinal hemorrhage, intracranial hemorrhage), followed by osteofascial compartment syndrome, renal failure, and even multiple organ failure. According to our experience, during VA-ECMO support, heparin was used as an anticoagulant, and active clotting time was maintained at 160s-180s. Plasma and platelets are transfused as early as possible due to the destruction of blood by ECMO and abnormal coagulation after surgery. After hemodynamic stabilization, ventilator-assisted respiration is discharged as early as possible, and enteral nutrition is administered, which avoids ventilator-associated pneumonia, reduces the risk of hemorrhage, and determines the recovery of neurological function in the postoperative period. The implications of this complication are devastating as the ELSO registry data reports that only 26% of ECMO patients who develop intracranial hemorrhage survive to discharge [7], so early cranial decompression once intracranial hemorrhage is diagnosed. In the past, we encountered a case of postoperative type A aortic dissection who developed cerebral hemorrhage, and the patient was cured after emergency craniotomy and decompression.

We have dealt with a patient with surgical repair of acute type A aortic dissection concomitant CABG and ECMO support, the patient developed osteofascial compartment syndrome in the lower limbs after surgery, we decompressed the patient through early incision, daily removal of some necrotic tissues, iodine-hydrogen peroxide-saline rinsing (Figure 2), and we put blood filters on ECMO, which is good for removing toxins in the blood and maintaining electrolyte stabilization at a later stage, through the implantation of the skin, the patient survived (Figure 3). In our series, the femoral artery served as an arterial cannulation site for the ECMO arterial cannula. Femoral cannulation is easier and faster to perform when the patient is suffering from an unstable hemodynamic condition. the cannula may also exert a so-called downstream compression effect, which limits the blood flow below its insertion point [8]. The femoral artery should be palpated before intubation to avoid severe calcification. A 5-0 polypropylene purse-string is then performed on the vessel. The purse-string should be in the longitudinal direction, and as small as possible, in order to avoid stenosis of the artery after cannula removal and purse-string knotting. Finally, a 10Fr sheath tube was inserted below the femoral artery cannula to provide lower limb blood flow. And Doppler ultrasonography (D-US) was used to monitor peak systolic velocity (PSV) of distal arteries, such as the posterior tibial or dorsalis pedis, generally, the flow velocity of the posterior tibial artery should be greater than 10cm/s for adults. Retrograde blood inflow from cannulas through femoral artery could lead to false lumen dilation and be not conducive to the recovery of heart function. Now we preferred to place arterial cannulas through axillary artery to provide antegrade perfusion (Figure4). But this method also carries the risk of osteofascial compartment syndrome as well as nerve damage resulting in upper extremity edema. Frenckner B et al reported axillary artery cannulation is, however, technically more difficult than the femoral artery and there is also a risk of hyper-perfusion of the ipsilateral arm and brain [9].

**Fig 2.**
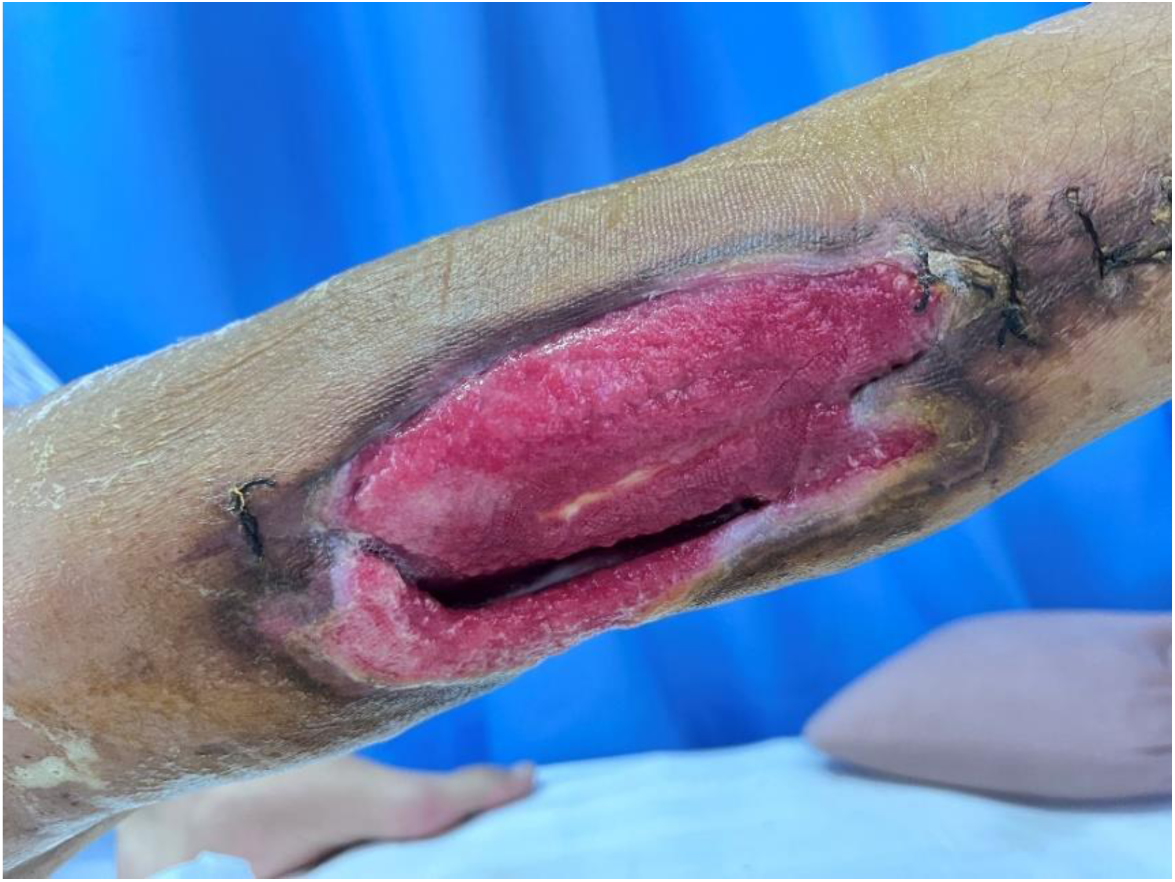
The incision of low limb osteofascial compartment syndrome.

**Fig 3.**
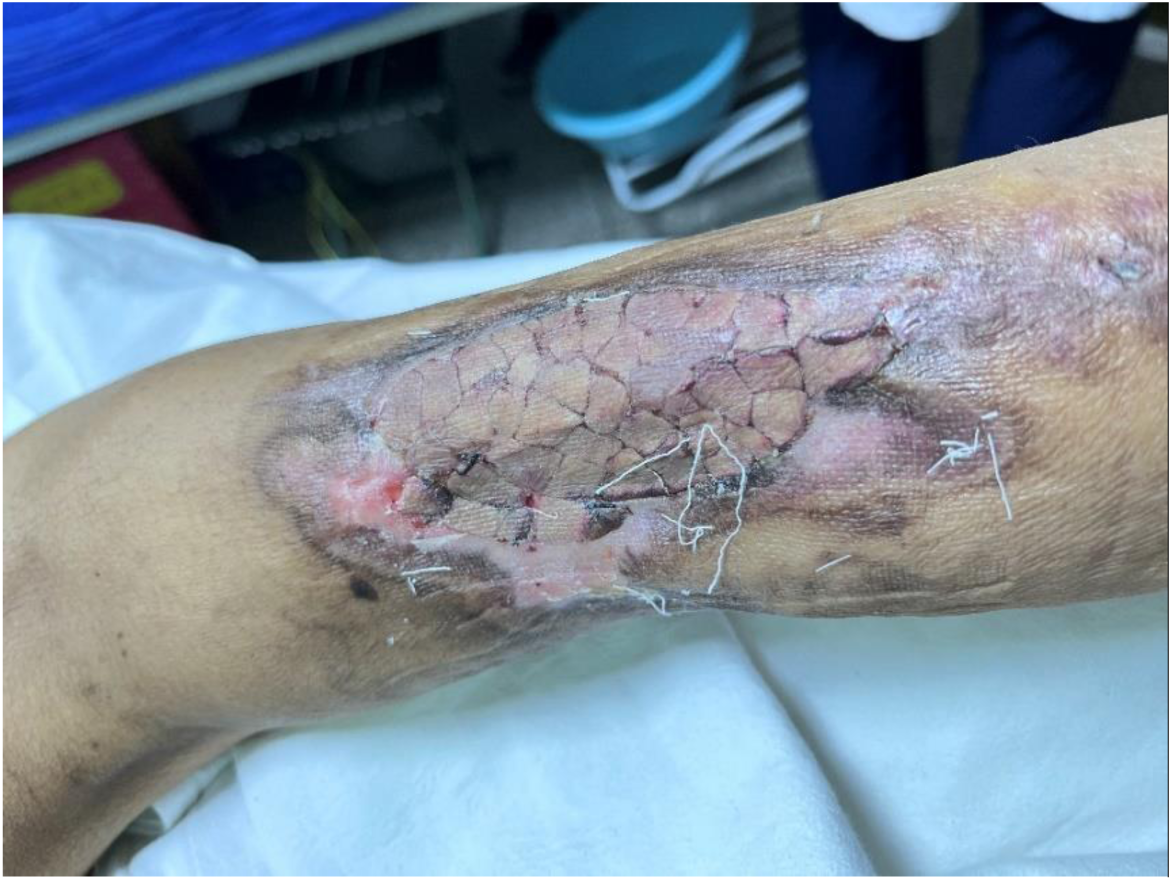
The skin grafting of low limb osteofascial compartment syndrome.

**Fig 4.**
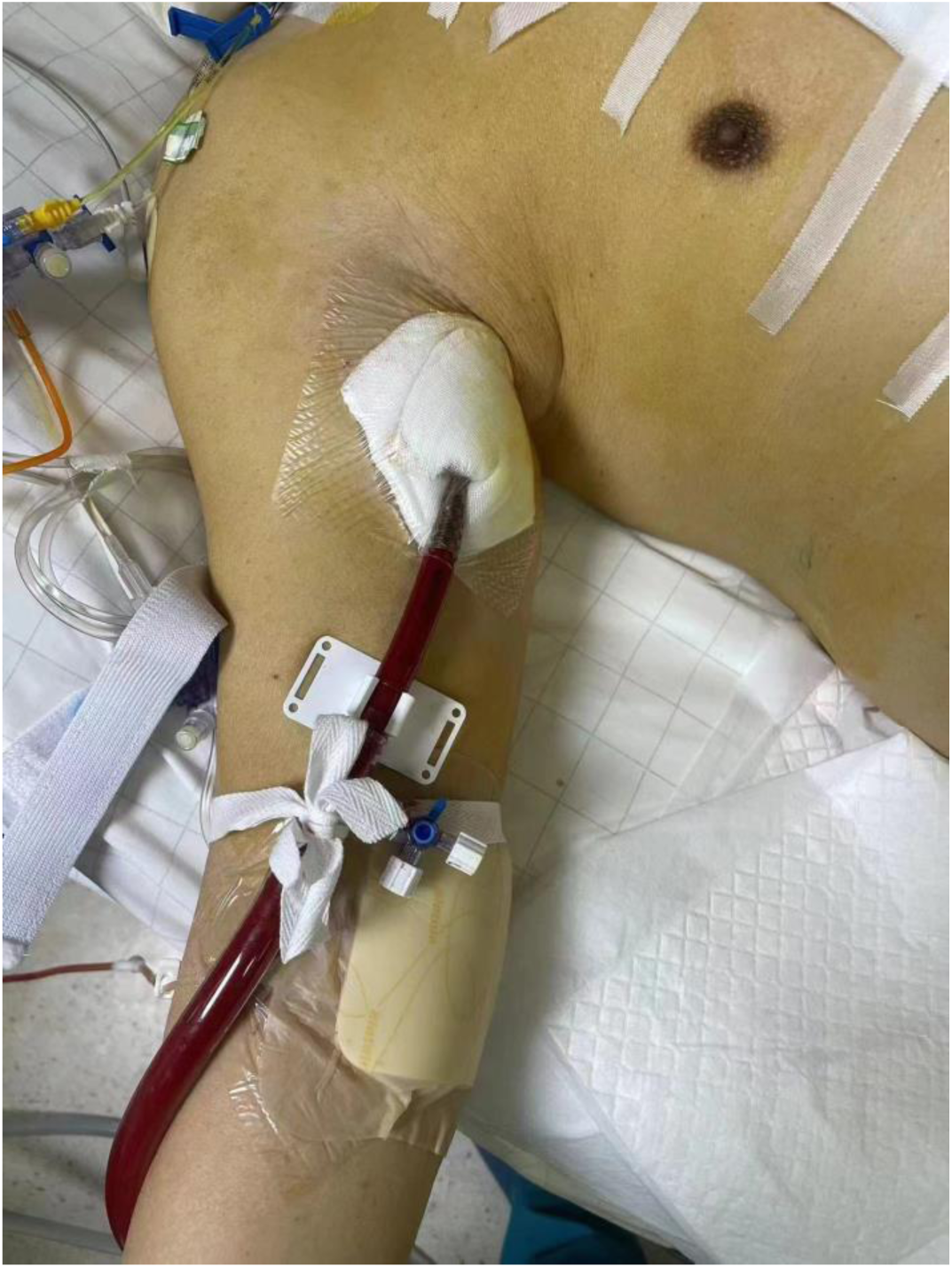
The arterial cannulas through axillary artery.

Nevertheless, our study was a retrospective, single-center study. And the population of TAAD patients who need CABG or ECMO support remained relatively low and bias might exist influencing the homogeneity of the study. In the future, we will consider multivariate regression, propensity matching analysis and multicenter controlled research may provide more evidence to support our data. Finally, with the advances in surgical techniques, anesthesia, and perioperative care, they surely affect the surgical results of different periods.

## Conclusions

In conclusion, in our study, for the patients who failed to wean from CPB, surgical repair of acute type A aortic dissection Concomitant CABG can provide more excellent short and midterm outcomes than ECMO support. However, concomitant CABG are also associated with long-term complications of the great saphenous vein embolization and severe tricuspid valve regurgitation.

## Data Availability

The data that support the findings of this study are available from the corresponding author upon reasonable request.

## Declarations

### Ethics approval and consent to participate

This observational study was approved by the Ethics committee of Yangzhou University Affiliated Northern Jiangsu People’s Hospital. Written informed consent had been obtained from each participant.

### Consent for publication

We hereby grant consent for the publication of this work.

### Authors’ contributions Conceptualization

XY.W, XL.W; Supervision: MJ.G, Z.Y, B.L, XP.C; Medical writing–draft, review & editing: D.Z, GJ.Z, YS.S. All authors have read and agreed to the latest version of the manuscript.

## Acknowledgements

Thanks to Director Xing-Peng Chen and Director Yu-Sheng Shu for their guidance and help.

## References

[1] Evangelista A, Isselbacher EM, Bossone E, Gleason TG, Eusanio MD, Sechtem U, et al. Insights from the International Registry of Acute Aortic Dissection: a 20-Year Experience of Collaborative Clinical Research. Circulation. 2018,137:1846–1860.

[2] Shimizu H, Okada M, Tangoku A, et al. Thoracic and cardiovascular surgeries in Japan during 2017: annual report by the Japanese Association for Thoracic Surgery. Gen Thorac Cardiovasc Surg. 2020,68:414–449.

[3] Jun-Neng Roan, Hsuan-Yin Wu, Luo CY. Risk factor analysis of surgical and long-term results in patients with acute type A aortic dissection. Recent advances in acute type A aortic dissection. 2015,265–285.

[4] Kai Zhang, Song-Bo Dong, Xu-Dong Pan, et al. Concomitant coronary artery bypass grafting during surgical repair of acute type A aortic dissection affects operative mortality rather than midterm mortality. Asian Journal of Surgery. 2021,44:945–951.

[5] Raffa GM, Kowalewski M, Brodie D, et al. Meta-Analysis of Peripheral or Central Extracorporeal Membrane Oxygenation in Postcardiotomy and Non-Postcardiotomy Shock. Ann Thorac Surg. 2019,107(1):311–321

[6] Okada K, Omura A, Kano H, et al. Recent advancements of total aortic arch replacement. J Thorac Cardiovasc Surg. 2012,144:139–145.

[7] Lorusso R, Gelsomino S, Parise O, et al. Neurologic injury in adults supported with veno-venous extracorporeal membrane oxygenation for respiratory failure: findings from the extracorporeal life support organization database. Crit Care Med. 2017,45:1389–1397.

[8] Eleonora B, Gennaro M, Jorik S, et al. Limb ischemia in peripheral veno-arterial extracorporeal membrane oxygenation: a narrative review of incidence, prevention, monitoring, and treatment. Crit Care. 2019,23(1):266.

[9] Frenckner B, Broman M, Broome M. Position of draining venous cannula in extracorporeal membrane oxygenation for respiratory and respiratory/circulatory support in adult patients. Crit Care. 2018,22:163.

